# Pro-ictal, rather than pre-ictal, brain state marked by global critical slowing and local gamma power increase

**DOI:** 10.1101/2024.10.28.24316105

**Authors:** I. Dallmer-Zerbe, J. Kopal, A. Pidnebesna, J. Curot, M. Denuelle, A. De Barros, J.C. Sol, L. Valton, E.J. Barbeau, J. Hlinka

## Abstract

**Objective:** The pre-surgical evaluation of epilepsy relies on the electrophysiological recordings of spontaneous seizures. During this period drug dose decreases increase the likelihood of seizures transitioning the brain from a low to high seizure likelihood state, so-called *pro-ictal* state. This study aimed to identify the dynamic brain changes characteristic of this transition from 386 ten-minute segments of intracranial EEG recordings of 29 patients with drug-refractory temporal lobe epilepsy.

**Methods:** We studied brain dynamics through mean phase locking value and relative power in gamma band, and autocorrelation function width. We further explored interactions with pro-ictal factors, such as rate of interictal spikes and high frequency oscillations, circadian and multi-day cycles, and clinical outcomes.

**Results:** We observed significant increases in gamma power in the epileptogenic zone, and critical slowing in both the epileptogenic zone and presumably healthy cortex. These changes were linked with increases in spike and high frequency oscillations rate.

**Conclusions:** Brain dynamics changed on the slow time scale - from the beginning to the end of the multi-day interval - but did not change in the short-term during the pre-ictal interval, thus could reflect pro-ictal changes.

**Significance:** We highlight gamma power and critical slowing indices as markers of pro-ictal brain states, as well as their potential to track the seizure-related brain mechanisms during the presurgical evaluation of epilepsy patients.

**Highlights:**

- Study of multi-day brain dynamics during presurgical evaluation in drug-resistant temporal lobe epilepsy.
- Increasing seizure susceptibility is marked by a gamma power rise in the epileptogenic zone and network-wide critical slowing.
- Changes occur over days, not minutes before seizures, suggesting a multi-day pro-ictal phenomenon during presurgical evaluation.

## 1. INTRODUCTION

Ictogenesis, as often studied in the context of seizure prediction, is a complex process (Jiruska et al., 2010). It is typically studied on a time scale of minutes up to hours before seizure (Mormann et al., 2007). Previous studies have linked the time period before seizure, i.e. the *pre-ictal state*, to be characterized by a progressive and global increase in neuronal activity, a build-up of low-amplitude high-frequency activity (>100 Hz) and a reduction in system complexity (Jiruska et al., 2010). Immediately before seizure onset, the brain is in a highly sensitive state where even weak perturbations can initiate seizures (Chang et al., 2018; Jiruska et al., 2010). Among the notable changes observed in iEEG recordings in this time interval is critical slowing down (Chang et al., 2018; Lepeu et al., 2024; Maturana et al., 2020). Critical slowing refers to a phenomenon where a system takes longer to recover from small perturbations, indicating an increased risk of transitioning to a different state, such as a seizure. Other notable iEEG changes during ictal transitions as evidenced by several studies are increases in the rate of interictal epileptic discharges, or spikes, and high-frequency oscillations (HFOs > 80 Hz) (Jacobs et al., 2009; Karoly et al., 2016; Malinowska et al., 2015; Scott et al., 2021), as well as aberrant gamma (30-100 Hz) dynamics (Hughes, 2008; Janca et al., 2021; Medvedev et al., 2011; Meisel et al., 2015; Mormann et al., 2000; Weiss et al., 2015; Zweiphenning et al., 2019). Such aberrant dynamics are often found within the epileptogenic network (Bartolomei et al., 2017), and particularly in the EZ (Medvedev et al., 2011; Sumsky and Greenfield, 2022; Weiss et al., 2015) outside of the epileptogenic network (Naftulin et al., 2018).

However, the reliable mapping of pre-ictal states remains a challenge (Freestone et al., 2015; Kuhlmann et al., 2018; Mormann et al., 2007; Mormann and Andrzejak, 2016; Usman et al., 2019; Wilkat et al., 2019). Overcoming it requires overcoming dataset limitations (Kuhlmann et al., 2018; Stacey et al., 2011) and achieving better understanding of the brain dynamics at play, as well as their influencing factors. Accordingly, epilepsy research has recently shifted its focus from the short-term (and deterministic) prediction of single seizure events to the long-term (and probabilistic) prediction of time intervals of increased seizure risk, searching for so-called *pro-ictal states and their* markers, that act on timescales of days, months and even years (Baud et al., 2020). When taking into account these slow fluctuations in seizure likelihood, seizure prediction becomes seizure forecasting (Baud et al., 2018; Cook et al., 2013; Freestone et al., 2015; Karoly et al., 2018; Proix et al., 2021; Stirling et al., 2021). Examples of common observations in epilepsy linked to these fluctuations are seizure clustering (several seizures occurring close together in time) and the variations in brain responses to the same stimulation at different time points (Chang et al., 2018; Jiruska et al., 2010; Kudlacek et al., 2021; Pérez-Cervera and Hlinka, 2021). Similarly suggesting that seizures are modulated by slow dynamics, seizures closer together in time were reported to be more similar (Schroeder et al., 2020), and seizures were shown to be influenced by previous seizures (Kudlacek et al., 2021). Moreover, a growing body of studies have reported pro-ictal factors that influence seizure likelihood (Bernard et al., 2023; Karoly et al., 2016; Panagiotopoulou et al., 2022; Payne et al., 2021), such as patient-specific sleep-and-wake cycles (Ferastraoaru et al., 2018; Karoly et al., 2018), administered drug dose, weather, and temporal factors (e.g. time of day, day of week, or lunar phase) (Bernard et al., 2023; Meisel, 2020; Meisel et al., 2015; Payne et al., 2021). However, recent evidence also questions the influence of external multidien cycles on seizure occurrence (Leguia et al., 2021; Rao & Leguia et al., 2021). Thus, the complex link between the epileptic brain processes and such pro-ictal factors remains insufficiently understood (Rao & Leguia et al., 2021).

Combining tools from seizure prediction and forecasting, this study aimed to investigate the multi-day pre-seizure mechanisms during pre-surgical evaluation. We hypothesized a pre-ictal, short-term change in brain dynamics minutes immediately before seizure, linked with seizure initiation. Furthermore, we hypothesized there would be a progressive *pro-ictal, long-term* increase of seizure susceptibility in this multi-day interval linked with drug dosage decreases and other established pro-ictal factors. To test these hypotheses, we leveraged three *measures* of brain dynamics: one for gamma power alterations, the gamma power ratio (gPR) (Panagiotopoulou et al., 2022), one for gamma synchronization, the phase locking value in the gamma band (gPLV) (Bruña et al., 2018; Mormann et al., 2000), and one for critical slowing, the autocorrelation function width (ACFW) (Maturana et al., 2020). Furthermore, we studied their interaction with several pro-ictal *factors*: linear time (time since first recording), daytime, drug dosage, spike and high frequency oscillation (HFO) rate as well as signal-to-noise ratio (SNR) as a control variable to assess data quality. We compared pre-seizure dynamics between the EZ and healthy brain areas in the short- and long-term, i.e., investigating the measures’ changes firstly within the last minutes before the first seizure (short-term changes), and secondly from the beginning of the recording up until this first seizure (multi-day changes).

## 2. METHODS

### 2.1. iEEG Data acquisition

Our dataset consisted of intracranial EEG data recorded for 29 patients with drug-resistant epilepsy (Table 1). The patients were monitored at the Brain electrophysiology, Epilepsy and Sleep Unit, department of Neurology, at Toulouse University Hospital to identify and possibly resect the brain areas involved in seizure generation, between 2009 and 2020. If surgery was offered, its outcome was rated using the International League Against Epilepsy (ILAE) rating scale (Wieser et al., 2001). iEEG were retrospectively collected with the following inclusion criteria: over 12 years of age, temporal involvement in the epileptogenic network, recording electrodes implanted in the temporal lobe, and an available iEEG recording of the last 10 min before the first recorded seizure. Each patient received detailed information about the objectives of the SEEG technique before intracerebral electrode implantation. Data were collected using the computerized patient records of the Toulouse University Hospital IT system. This data collection was authorized in accordance with the French Data Protection Authority MR-004 reference methodology. Following assessment and validation by the Data Protection Officer and in compliance with general data protection regulations, the research, named Predire (as in Seizure Prediction), is registered in the official retrospective studies register of the Toulouse University Hospital, managed by the Research and Innovation Department (RnIPH 2020-130), and is covered by the MR-004 law (CNIL number: 22.6723 v 0). The ethical conditions of this study have been upheld and approved by the Toulouse University Hospital. Consent was obtained from all patients. The implantation was individually tailored to the hypotheses of EZ according to a noninvasive assessment, and the placement of each depth electrode was based exclusively on clinical criteria independently of this study. Standard intracranial electrodes (Microdeep depth electrode, DIXI medical, France) with 8-15 platinum/iridium contacts (diameter 0.8 mm, contact length 2 mm) were used. Intracranial EEG activity was recorded using two synchronized 64-channel acquisition systems (SystemPlus Evolution, SD LTM 64 EXgPRESS, Micromed, France) with varying sampling frequencies (at least 256 Hz) and high pass-filtered to 0.008 or 0.15 Hz. An example recording is given in Fig. 1A. Electrode placement across all patients covered a wide-spread brain network (Fig. 1B).

**Figure 1.**
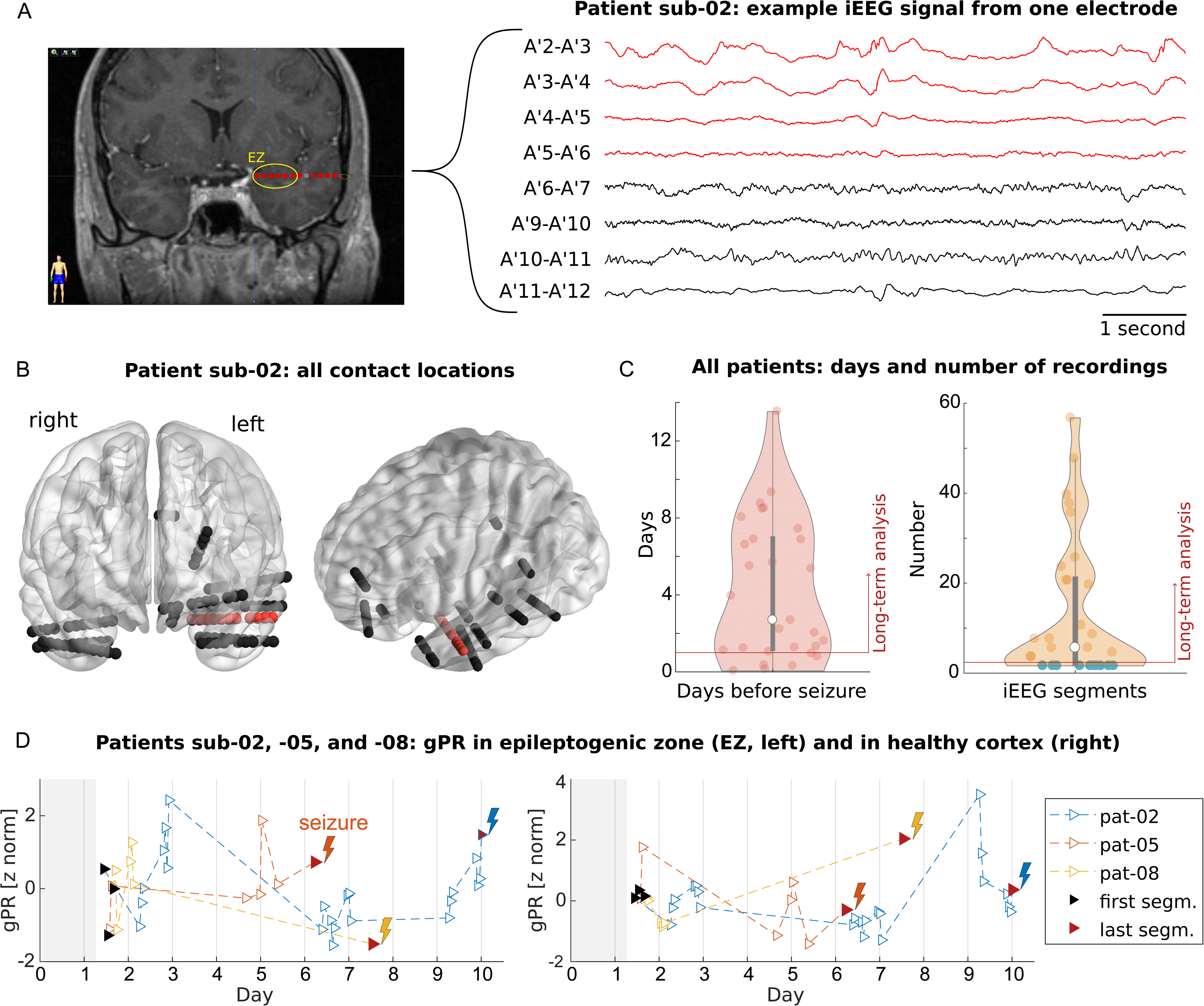
Data description and study design. **A:** Example iEEG of recording during interictal phase. The MRI scan on the left side shows an intracranial electrode targeting the left hippocampus in sub-02. Electrodes channels (macro contacts) in the epileptogenic zone (EZ) are highlighted in red. Schematic representation on the right side depicts example signals from the eight bipolar channels. Red traces correspond to bipolar channels located in EZ. **B:** Locations of all bipolar channels for the selected patient referenced in the MNI space ([BrainNet Viewer](https://journals.plos.org/plosone/article?id=10.1371/journal.pone.0068910)). The intracranial electrode targeting the left hippocampus is marked in red, **C:** Number of available recordings as well as days between electrode implantation and the occurrence of first seizure. Only patients with more than one day of recordings between the implantation (10 a.m. the second day after the procedure) and first seizure were included in the long-term analysis (left violin plot, red line). All patients were included in the short-term analysis (analyses described in Methods). The number of analyzed iEEG segments varied (mean = 13, std = 16) across the 29 participants (right violin plot). In the short-term analysis, only a single recording (in turquoise) was used per patient. **D:** Study setup exemplified in three patients: sub-02, sub-05, and sub-08 (see Table 1). Patients had different numbers of segments available (triangles), variably scattered across the interval between the day of their implantation (day 0) and the day of the seizure (here, day 6, 7 and 10). Recordings before 10 a.m. on day 1 were excluded to avoid the immediate effects of implantation (interval marked in gray). Mean measure (here gamma power ratio gPR) across channels in each recording was compared between different brain areas (left vs. right panel). Long-term changes were assessed by comparing only the first and the last recorded segment (black and red triangles). Short-term changes were assessed within the last recorded segment (red triangle). The lightning symbol indicates a seizure for a given patient.

**Table 1:**
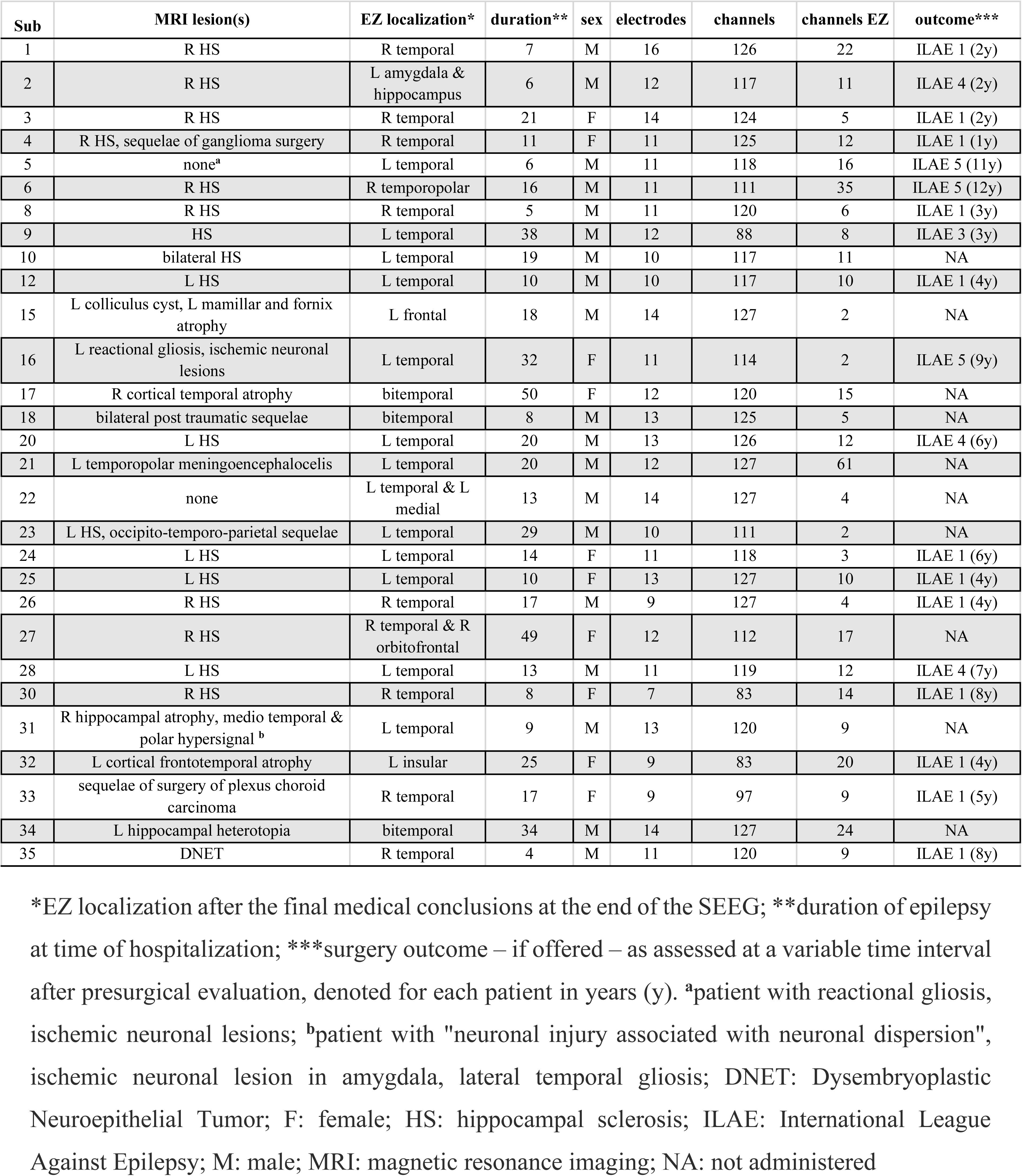
Patient Characteristics.

The determination of the different zones of the epileptic network was carried out by clinicians independently and blinded to our analyses. The epileptogenic zone is an electroclinical concept that corresponds to the network of brain regions responsible for generating seizures and should encompass the seizure-onset zone (Bartolomei 2017). To determine the epileptogenic zone, clinicians rely on the observation of the seizure onset pattern in intracranial EEG recordings (Lagarde et al. 2019, Abdallah et al. 2024). The contacts initially involved in this pattern define the epileptogenic zone. The irritative zone (IZ) corresponds to the site of interictal epileptic discharges, including interictal spikes and high-frequency oscillations (HFOs). However, IZ regions that did not overlap with the EZ were excluded from our analyses. The lesion zone is defined based on brain MRI as the area affected by a visible lesion. Healthy cortex is defined as regions with no overlap with the EZ, IZ, or lesion zone: there are no interictal epileptic discharges in the healthy cortex.

The iEEG recording started in the hours after electrode implantation and lasted the whole hospital stay. Due to storage limitations, the hospital did not keep all iEEG recordings. This study focused on the interval between implantation and first seizure, which was of variable length across patients (Fig. 1C). Furthermore, we only considered recordings from 10 a.m. the day after the surgical procedure in order to avoid the influence of anesthetics on the measured brain activity. If the interval between the first considered recording and first spontaneous seizure was less than a day, the patients were not considered for the long-term analysis (n = 19), but only the short-term analysis (n = 29; analyses described below). The number of resulting iEEG segments in the chosen interval varied among patients (Fig. 1D; mean = 13, std = 16 across all 29 patients; mean = 20, std = 16 across the 19 patients of the long-term analysis). The time points of recordings were scattered across the chosen interval (Fig. 1E), leaving gaps of up to a couple of days in between them (e.g., sub-08 in Fig. 1E).

### 2.2. Data processing

All processing of iEEG data was conducted with Matlab (Version 9.8.0, The Mathworks Inc, Natick, MA, USA) and the interactive Matlab toolbox EEGLAB (Delorme and Makeig, 2004). For each patient, we collected all available iEEG recordings before the occurrence of a first seizure (Fig. 1E). For each recording, we then extracted one 10-minute segment for every hour (always the first 10 minutes). As an example, an iEEG recording that was three-hour-long would be segmented into three segments.

Further preprocessing steps were applied to all iEEG segments. These steps included downsampling to 256 Hz, high pass filtering with a cut-off frequency of 0.5 Hz, removing non-EEG channels, re-referencing to bipolar montage (single bipolar montage is referred to as a “channel” throughout this study), and finally, the exclusion of error channels. For re-referencing, the difference in activity between (only) directly neighboring channels was calculated. Error channel rejection consisted of a two-step procedure using EEGLAB *pop_rejchan()* function with the ‘spec’ rejection method. Firstly, channels with significantly high power in the 48-52 Hz frequency range (>3 standard deviations from mean channel power in this range), and secondly, outlier channels with regards to the whole 1-128 Hz frequency range (>4 standard deviations from mean channel power) were identified. To obtain a consistent number of channels for each patient across all respective recordings, the same error channels (accumulated error channels over all segments) were excluded from all segments for a given patient. For the same reason, iEEG channels that were present in some but not all segments were disregarded.

### 2.3. Mapping brain dynamics

The aim of the presented study was to map the multi-day pre-seizure dynamics during presurgical evaluation. To explore the brain dynamics preceding the first spontaneous seizure, we studied the temporal evolution of brain behavior using measures of critical slowing, altered synchrony and altered gamma power (30-100 Hz) in all patients. Specifically, we calculated gamma power ratio (gPR; Fig. 1E), phase locking value in gamma band (gPLV), and autocorrelation function width (ACFW). To obtain robust measure estimates over the whole 10 min iEEG segment, we calculated the respective measures in 10 consecutive windows of 60 seconds lengths and averaged them across all windows. This approach was based on exploratory results of measuring stability in a two-hour-long uninterrupted iEEG recording from healthy cortex (Sup. Fig. 1).

#### 2.3.1. Power ratio in the gamma frequency band (gPR)

In order to provide insights into the dynamics behind neuronal synchronization, we estimated the power ratio in the gamma frequency band using Welch’s power spectral density algorithm for every 60s segment of the iEEG recording (Panagiotopoulou et al., 2022). Specifically, we computed power spectral density using a sliding window of 500 ms length and a shift of 16 ms. Each sliding window was multiplied by a Hanning window to smoothly taper the endpoints to zero and mitigate the discontinuity that produces leakage. The modified periodograms were averaged to obtain frequency-resolved power spectral density estimates for the 60s iEEG segment. Then, the vector of frequency-varying power spectral density estimates was log-transformed and subsequently Sigmoid transformed (*S*(*x*) = 1[1 + *exp*(−*x*)]^−1^) to ensure positive entries between 0 and 1 as in previous research on iEEG (Panagiotopoulou et al., 2022). As the last preprocessing step, we averaged (across segments) the preprocessed estimates for each of the five main frequency bands: *δ*: 1–4 Hz, *θ*: 4–8 Hz, *α*: 8–13 Hz, *β*: 13–30 Hz and *γ*: 30–100 Hz. Finally, the power spectral density estimate in the gamma band was normalized by the average power spectral density in all other bands to obtain the power ratio in the gamma frequency band.

#### 2.3.2. Phase locking value in the gamma band (gPLV)

Phase locking value (PLV) is a well-established measure of brain connectivity, especially in the case of signals such as M/EEG. PLV quantifies frequency-specific phase synchronization between two neuro-electric signals (Lachaux et al., 1999). In this study, we used an implementation of PLV from Bruña and colleagues (Bruña et al., 2018). It is important to note that the PLV implementation used in our study corresponds to the mean phase coherence introduced by Mormann et al. (2000). However, we chose to refer to it as PLV to avoid confusion, as our analysis focuses solely on phase relationships without incorporating amplitude.

For each 60-second window, we first applied a zero-phase FIR least-squares bandpass filter with a 15% transition width to isolate the gamma band (30–100 Hz) in each pair of iEEG channels. Next, we applied the Hilbert transform to extract the instantaneous phase from the analytic signal. Finally, we computed PLV to quantify phase synchronization in the gamma band (gPLV) between each pair of iEEG channels as:

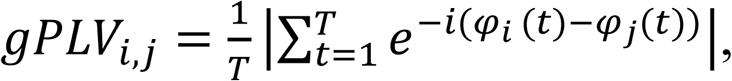

where T is the length of the iEEG signal, φ_i_ is an instantaneous phase as obtained by Hilbert transform of the iEEG signal and i, j correspond to the pair of channels for which gPLV is computed.

Resulting gPLV between all pairs of channels is represented by a matrix of real-valued numbers between 0 and 1.

#### 2.3.3. Critical slowing down: Autocorrelation function width (ACFW)

The change from normal to a seizing state can be viewed as a critical transition happening in the complex dynamical system (i.e., the brain). This system approaching the critical transition can be characterized by critical slowing down features, where the ACFW represents one of the most commonly used measures (Maturana et al., 2020). The autocorrelation function measures the correlation of the signal with its shifted copy. High autocorrelation for higher lags between the signal and its shifted copy indicates a smooth signal, while white noise, for example, has low autocorrelation for all positive lags. In this study, ACFW was estimated based on the implementation of Maturana and colleagues (Maturana et al., 2020); as the width at the half maximum of the autocorrelation function. Autocorrelation width yields estimates comparable to the lag-1 autocorrelation measure (Milanowski and Suffczynski, 2016) while offering the advantage of a larger dynamic range (Maturana et al., 2020). An increase in this measure would indicate a broadening of the autocorrelation function and thus a slowing of the observed dynamics, which might indicate that the system approaches the critical transition (seizure).

### 2.4. Pro-ictal factors

We further explore the relationship of estimated brain dynamics measures with other *factors* that could potentially explain changes in brain dynamics. Namely, we quantified the influence of administered drug dose, elapsed time from electrode implantation, time of the day, signal-to-noise ratio of iEEG recordings, as well as spikes and high-frequency oscillations present in the recordings. For each of these five *factors*, we derived a single metric to characterize the 10-minute iEEG segment.

#### 2.4.1. Drug dose

The factor of drug dose can reflect a patient’s seizure susceptibility. To facilitate seizure occurrence, anti-seizure medication was gradually reduced from the first day of recordings. Since each patient received different drugs and dosages, we normalized administered drug doses for comparability. Specifically, for each drug and for each patient, the actual dose was divided by the maximum dose given between admission and the first seizure. For a given patient and at each time point, the dosage was calculated as the mean of relative drug doses, averaged across all drugs administered at the most recent time of medication (e.g., drugs given at 6 p.m. were combined into a single score representing the medication level until the next administration at 9 p.m.). Since the resulting summary index ranges between 0 and 1, it is comparable across patients. In this mapping, the highest relative dosage (i.e., 1) corresponds to the lowest susceptibility to having a seizure.

#### 2.4.2. Linear Time

Seizure susceptibility was reported to fluctuate over daily timescales (Schroeder et al., 2020). We thus characterized each iEEG segment by the time elapsed from the iEEG electrode implantation.

#### 2.4.3. Daytime

Prior research shows that circadian timescales influence seizure susceptibility (Baud et al., 2018). To take into consideration the influence of the time of the day (i.e., day and night cycles) on the estimated measure of brain dynamics, we characterized each iEEG recording by daytime at the beginning of the recording, in 24-hour format (i.e. 14:02:35).

#### 2.4.4. Spikes and high-frequency oscillations (HFOs)

Spikes and HFOs were proven to be valuable biomarkers of epileptogenic tissues. In particular, the combination of both was shown to have superior prediction performance compared to individual measures (Roehri et al., 2018). We used the AnyWave: Delphos software (Colombet et al., 2015) to detect spikes and HFO (80-126 Hz; maximum frequency limited by the 256 Hz sampling frequency) in each 10 min iEEG segment separately for each channel. Each channel was then characterized by the geometric mean between spike and HFO rate:

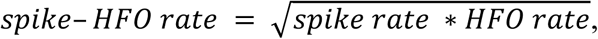

where the spike rate corresponds to a per-minute estimate of the number of spikes in each channel (identically for HFO rate). We used mean spike and HFO rate across all channels to represent the amount of spikes and HFOs present in a single recording (Roehri et al., 2018). Using geometric means ensures that the spike and HFO rate is sensitive to both spikes and HFOs, emphasizing their co-occurrence. Physiologically, a high spike and HFO rate indicates strong synchronization between spikes and HFOs, which is a hallmark of epileptogenic tissue. In contrast, a low spike or HFO rate suggests a reduced spike rate and / or HFO rate. This reduced co-occurrence is more characteristic of healthy regions, where spikes and HFOs occur independently rather than as a coupled pathological process.

#### 2.4.5. Signal-to-Noise-Ratio

As the last factor we quantified the signal-to-noise ratio (SNR). We use SNR as a control assessment that quantifies the clarity and strength of the neural activity. We quantified SNR in each iEEG segment for each channel separately. The used *snr* function from Matlab’s Signal processing toolbox returns the signal-to-noise of the input iEEG signal by comparing the power of the signal estimated from power spectral density to the power of the noise. In this context, noise is defined as the signal content that remains after excluding the power of the fundamental frequency and its first five harmonics, calculated using a modified periodogram. SNR is estimated across the entire frequency spectrum of the signal, providing a comprehensive assessment of signal quality. The resulting SNR value is in decibels (dB).

### 2.5. Description of the data analyses

#### 2.5.1. Short-term changes, trend analysis

The examination of short-term changes was based on the analysis of trends which are represented by the correlation of the minute-resolved measures (gPR, gPLV, ACFW) in the last 10 minutes before seizure onset (so-called “Last” segment), with time. We tested whether the trends in EZ are higher than those in healthy cortex using the Wilcoxon signed rank test across patients, at 0.05 significance level.

#### 2.5.2. Long-term changes, first-to-last

To assess long-term changes in brain dynamics, we compared the “First” iEEG recording segment after 10 a.m. the day after electrode implantation (to avoid immediate effects of the implantation procedure, such as effects of narcosis medication) with the “Last” iEEG recording segment right before the seizure. Specifically, we compared the measures characterizing the first segment to the measures of the last segment using the Wilcoxon signed-rank test. Subsequently, we tested whether the change (Last - First) in EZ was greater than the change in healthy cortex (defined as channels remaining unlabeled by clinicians; thus, excluding also other zones, such as irritative and lesion zones) using Wilcoxon signed rank test at 0.05 significance level (i.e., Change in EZ - Change in healthy cortex > 0). All resulting p-values were FDR-corrected using the Benjamini/Hochberg procedure across the three measures (and the two zones for the first comparison).

In a post-hoc exploration, we quantified the association strength of the measure changes with changes in the *factors* and continuous clinical variables (age, time since epilepsy diagnosis at the time of hospitalization, and the number of electrodes in EZ) using Spearman’s correlation. Finally, we used a Wilcoxon signed rank test to examine the relationship between measure change and binary clinical variables (sex, hemisphere of EZ, resection hemisphere, surgery outcome), at 0.05 significance level.

#### 2.5.3. Long-term changes, dissimilarity

Our final objective was to investigate brain dynamics using all available iEEG recordings, rather than limiting the analysis to only the First and Last segments as done previously. This approach allows us to capture a more continuous representation of how brain activity evolves over time. Given the irregular sampling of iEEG recordings across patients, a direct comparison of sequential time points was not feasible. To address this, we adopted the methodology proposed by (Schroeder et al., 2020), which enables a pairwise comparison of all available iEEG segments within each patient.

Specifically, as described previously, each patient’s brain dynamics were characterized by a measure (e.g., gPR) computed over 10-minute iEEG segments for each channel. Thus each iEEG segment was represented by a brain dynamics vector, with dimensions (number of channels × 1), where each entry represents the computed measure for a specific channel at that time point. To assess how brain dynamics change across time, we quantified the dissimilarity between all pairs of iEEG segments. This was achieved by computing the mean L1 distance between their corresponding brain dynamics vectors, capturing differences in brain activity patterns across channels. The result was a patient-specific pairwise distance matrix, where each entry represents the degree of dissimilarity between two iEEG segments recorded at different times.

To investigate how brain dynamics relate to our five factors, we quantified the dissimilarity of linear time, daytime, drug dose, and spike and HFO profiles. Linear time and daytime dissimilarities were computed based on the time elapsed between two iEEG recordings. Drug dose dissimilarity was calculated as the absolute difference between the relative medication doses at two time points. Spike and HFO profile dissimilarities were determined by computing the L1 distance between their respective spike and HFO rates. An identical procedure was applied to signal-to-noise ratio dissimilarities. By computing these factor dissimilarities, we aimed to systematically evaluate whether and how variations in external influences (time and medication) and intrinsic epileptiform activity (spike and HFO profiles) relate to shifts in brain dynamics over time.

Finally, we assessed the relationship between brain dynamics dissimilarities and factor dissimilarities by computing Spearman’s correlation between the upper triangular elements of the brain dynamics dissimilarity matrix and those of each factor dissimilarity matrix.

## 3. RESULTS

### 3.1. iEEG dataset and patients’ characteristics

We studied multi-day intracranial EEG data from 29 patients with drug-resistant, temporal lobe epilepsy (see Table 1: Patient Characteristics). The patients’ mean age was 33 ± 13 (range: 12 to 58) years and mean epilepsy duration was 18 ± 12 (range: 4 to 50) years. The patient sample comprised 19 males and 10 females. Patients had on average 12 ± 2 (range: 7 to 16) implanted electrodes and 116 ± 13 (range: 83 to 127) recording channels. Out of those, EZ, as identified by the clinicians during monitoring, included on average 13 ± 12 (range: 2 to 61) channels, which were localized in the left hemisphere in 16, right in 10 and in both hemispheres in three patients. Surgery was later conducted in 19 of the patients, achieving seizure freedom in 12 patients (ILAE score 1) one year or more after surgery and improvement despite remaining seizures in 4 patients (ILAE score 2-4 (Wieser et al., 2001).

### 3.2. Short-term, pre-ictal changes in brain dynamics minutes prior to seizure

In the first step of the analysis, we focused on brain dynamics in the immediate time window around the onset of the first seizure (Fig. 2) that would be indicative of the incoming seizure. Specifically, we investigated whether there was a short-term increase in the brain measures within the last 10 minutes preceding seizure onset (i.e., in the Last segment, marked as red triangle in Fig. 1D). Therefore, we computed the three measures (gPR, gPLVMPC, ACFW) for every minute of this last segment. We then correlated the resulting measure values with time. These correlation coefficients we call trends. Positive trends indicate a linear increase in the measure before seizure onset, while negative trends indicate a linear decrease over time. We did not find any increasing trend on a group level (n = 29) in EZ compared to healthy cortex for either of the measures (ACFW: Z = 1.124, pFDR = .130; gPR: Z = −0.681, pFDR = .0.611; gPLVMPC: Z = 0.660, pFDR = .611).

**Figure 2.**
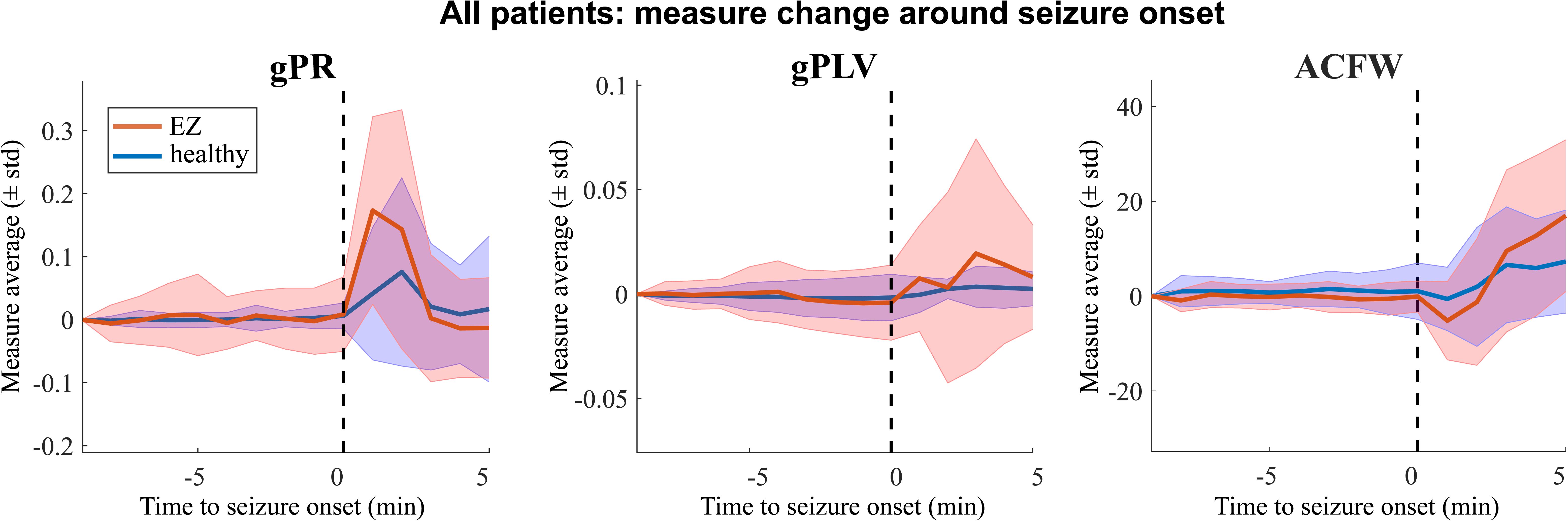
Short-term changes around seizure onset. Measure mean and standard deviation before and after the onset of the seizure (dashed line). After seizure onset, gPR and gPLV showed an overall increase, while ACFW displayed a decrease in EZ. Measure changes around the seizure onset in the healthy cortex were weaker compared to EZ. The solid line depicts the mean measure across all patients. The shaded area represents the standard deviation of the given measure.

### 3.3. Long-term, pro-ictal changes between electrode implantation and first seizure

We then investigated long-term changes in gPR, gPLV, and ACFW brain measures that could indicate a slow build-up in the brain mechanisms leading up to the first seizure during the interval of pre-surgical assessment. To that end, we compared the first iEEG recording segment after electrode implantation with the last iEEG recording segment before the seizure (red and black triangles in Fig. 1D). We included only patients in which this interval was at least one day (n=19). The change from the first to the last segment was thus computed as the difference: (Last - First), so that a positive change would indicate a long-term increase in the respective brain measure.

We first investigated whether the measures captured similar brain behavior. Thus, we quantified the relationship (across patients) between the changes of ACFW, gPLV, and gPR. Fig. 3A shows the Spearman correlation between any pair of measures. We did not observe any significant correlation, neither in EZ nor in the presumably healthy cortex. This absence of significant association indicates that the measures capture different aspects of brain activity.

**Figure 3.**
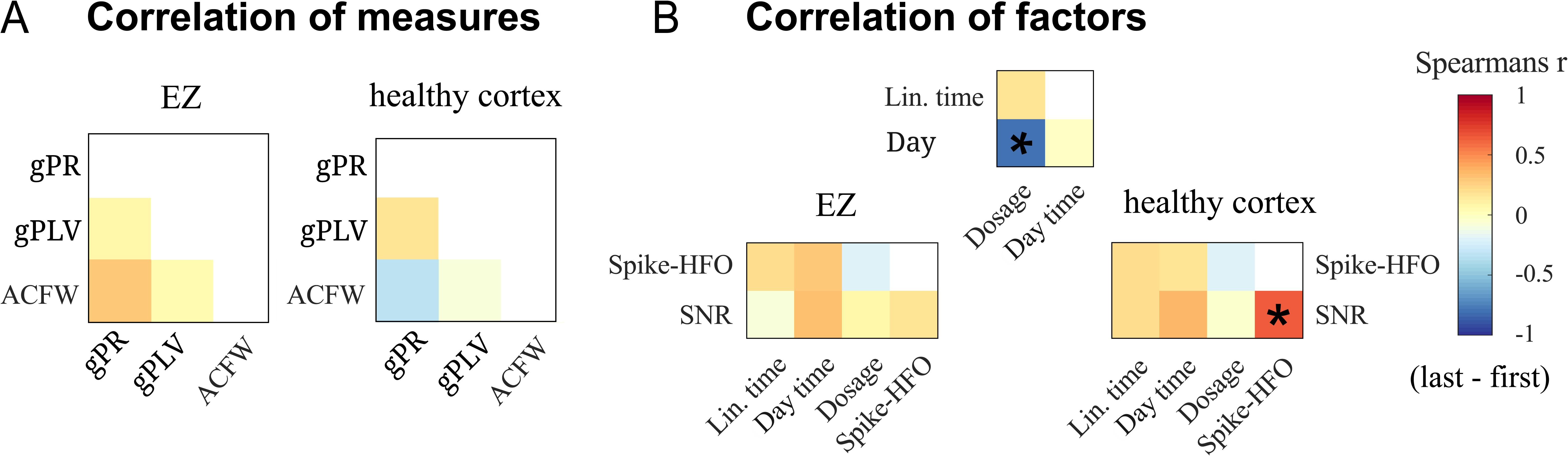
Measure and factor correlations between (Last - First) in EZ and healthy cortex. **A:** The measures did not correlate significantly one with the other, indicating that the measures capture different aspects of brain activity. **B**: The external factors (i.e., Day time, Dosage, Lin. time) showed expected correlations of drug dose with time. Furthermore, correlating external factors with EEG-based measures revealed significant correlation of spikes and HFOs with SNR (noise) in the healthy cortex.

We further repeated the same procedure to probe the interdependence of *factors*. Fig. 3B shows the correlation between any pair of *factors*. As one aspect of the clinical procedure is to reduce anti-seizure medication until occurrence of the first seizure, patients with a longer time until first seizure had a higher decrease in drug dose (*r* = −.800, *p* < .001). Furthermore, we did not expect spikes and HFO in the healthy cortex except for some false alarms detected by our automatic procedure. In accordance, the rate of spikes and HFOs correlated with the level of SNR in the healthy cortex (*r* = .647, *p* = .003). In summary, we confirmed our expectations regarding the interdependence of *factors* and demonstrated the independence of the brain measures under investigation.

After establishing the relationships between the measures, we studied the change in ACFW, gPLV and gPR from the first to the last iEEG recording. In line with a slow build-up in the pre-seizure brain dynamics, we expected to find a long-term increase in the measures localized to EZ. For statistical assessment, we tested for such an increase in EZ compared to the change in the healthy cortex separately for the three measures (FDR corrected to control for multiple comparisons). We found that most patients showed an increase of gPR, i.e, relative gamma power, in EZ but not in the healthy cortex (Fig. 4A, left panel). Accordingly, across patients there was a significant localized increase in gPR (one-sided, Wilcoxon signed rank test of Change in EZ - Change in CTRL > 0; *Z* = 2.515, *p_FDR_* = .033, Cohen’s *d* = .678). The change in gPLV, i.e. gamma synchrony, while pointing into the expected direction (Fig. 4A, middle panel), did not reach the significance level after FDR correction (*Z* = 1.912, *p_FDR_* = .077, *d* = .258). These results did not change when removing the two outlier patients. Finally, for ACFW (Fig. 4A, right panel), i.e. critical slowing, we did not find the expected localized increase in EZ as compared to CTRL (*Z* = −0.584, *p_FDR_* = .720, *d* = −.300). However, instead of a localized increase, we found a network-wide increase in ACFW with significant increases in both healthy cortex (*Z* = −2.757, *p_FDR_* = .009, *d* = −.680) and EZ (*Z* = −1.992, *p_FDR_* = .046, *d* = −.545), when tested separately. In summary, we found indications of a pro-ictal, long-term build-up in the brain dynamics during presurgical evaluation as captured by a localized increase of gamma power, but not synchrony, in EZ and a network-wide increase of the critical slowing index.

**Figure 4.**
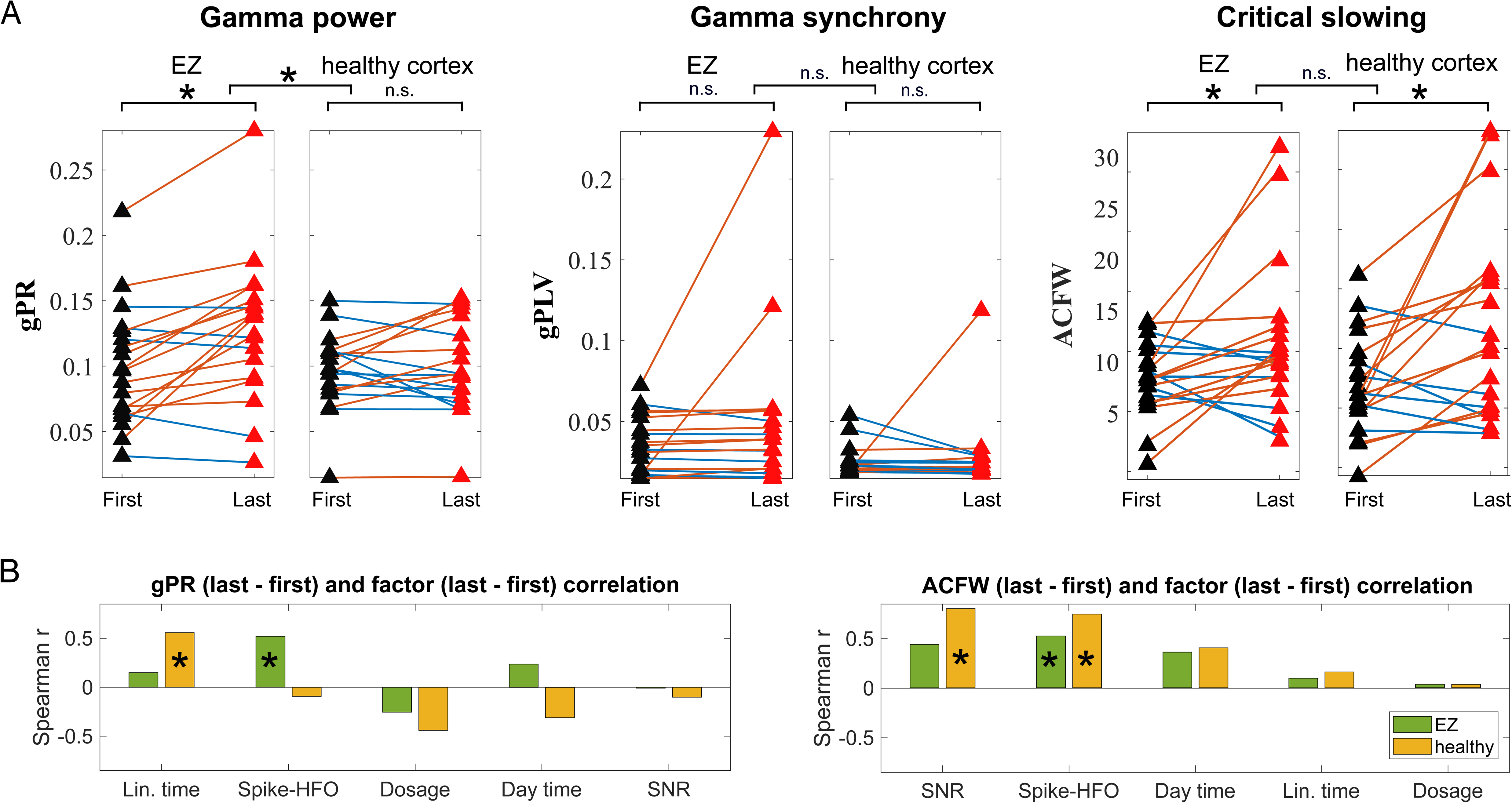
Long-term changes between the first and last segment. **A:** Changes of measures in EZ vs. healthy cortex for all patients. Relative gamma power (gPR: 30-100 Hz) in EZ of most patients displays an increase (red lines) as opposed to a decrease (blue lines). Gamma phase locking value (gPLV) change did not reach significance after FDR correction. Changes of autocorrelation function width (ACFW) are observed both in EZ and healthy cortex. **B:** Post-hoc analysis of gPR and ACFW change: correlation of first-to-last changes with the changes in factors. The gPR increase in EZ was accompanied by an increase in Spike and HFO rates. The ACFW increase in both healthy cortex and EZ was accompanied by an increase in Spike and HFO rates. The factors are plotted in descending order based on the correlation strength. n.s.: not significant; SNR: signal-to-noise ratio. * represents p < 0.05.

In a post-hoc exploration, we searched for the association between the gPR and ACFW increases and changes in the *factors* (Fig. 4B). In EZ the increase in gPR was significantly positively correlated with changes in spike and HFO rate (Spearman’s *r* = .521, *p* = .022). Thus, changes in gamma power in EZ from the beginning of a patient’s recording until immediately before their first seizure were associated with changes in spikes and HFOs. In the healthy cortex, as expected, we found no such relationship between gPR increase and spikes and HFOs. Similarly, the network-wide ACFW increase was accompanied by an increase in spikes and HFOs, both in the healthy cortex (*r* = .744, *p* = .000) and EZ (*r* = .523, *p* = .023). Thus, spikes and HFOs were consistently linked to the multi-day changes of brain dynamics during the multi-day pre-seizure interval. Other factors were less consistently linked. Notably, we did not find a significant relationship between the observed changes in brain dynamics and the time it took until seizure occurred or the changes in administered drug dose.

Finally, as a sensitivity analysis, we probed whether the identified gPR and ACFW increases were associated with clinical and other patient-specific variables (Sup. Fig. 2 - 5). Specifically, we tested the relationship between the measure increase with age and time since epilepsy diagnosis at the time of hospitalization (duration’ in Table 1: Patient Characteristics). We further tested for a group difference in the gPR increase based on sex (male vs. female), hemisphere of EZ and resection (left vs. right), and surgery outcome (ILAE 1 vs. ILAE 3-5). We did not find any significant association or difference between any clinical variable and the measure increases. Thus, the gamma power and autocorrelation increases can be interpreted as rather general phenomena in the context of (drug-resistant) temporal lobe epilepsy. When extending the analysis to further characteristics, such as number of recordings, electrodes in EZ and recordings during nighttime (Sup. Fig. 6), we found one significant correlation, which was between the numbers of electrodes in EZ and the change in ACFW measure change from first to last.

### 3.4. Pro-ictal brain dynamics are related with factor dynamics

Finally, we aimed to study the long-term dynamics of the observed changes in brain dynamics using all available iEEG recordings (compared to the above analysis of First and Last recordings only; see Fig. 1). To that end, we conducted a dissimilarity analysis between all pairs of iEEG segments extracted from all the available recording time points. We expected a gradual increase in gPR and ACFW during the whole-time interval, indicated by a positive correlation between the temporal distance of two data segments and their observed gPR difference. In addition, we speculated that pairs of segments recorded at a similar time of the day, a similar drug dose administered, or with a similar number of spikes and HFOs would have more similar gPR, indicating a relationship between long-term pre-seizure brain dynamics and the factors.

In this long-term analysis, for each patient with at least a day between the first segment and first seizure occurrence (n=19), we compared each available segment with all other segments, separately for the *measures* that had shown significant results in the previous analysis (gPR and ACFW) and the *factors* (linear time, daytime, drug dose, spike and HFO rate, SNR). In doing so, we obtained one dissimilarity matrix for each measure as well as for each factor per patient (Figure 5A, *top*). Within a given patient, we then correlated the dissimilarity matrices of gPR with those of the factors, resulting in one correlation value for each factor (Figure 5A, *bottom*). Based on the comparison of factor-specific correlation coefficients across patients, we observed similar factor interactions for EZ and healthy cortex (Figure 5B). On average, the dissimilarity of each factor correlated positively with the dissimilarity of gPR. In other words, across patients, pairs of segments that were more similar in their factor value (e.g. closer in time) were also more similar in gPR. Among the different *factors*, spikes and HFO rate showed the strongest link, while drug dosage showed the lowest similarity on average. On a patient level, the correlation coefficients varied (e.g. correlation of drug dose dissimilarity and gPR dissimilarity in EZ from *r* = −0.293 to *r* = 0.828). Thus, while in some patients, there were very strong indications of a gradual build up in pre-seizure brain dynamics, circadian time- and/or drug-dose dependence, in others, such indications were weaker. Overall, the dissimilarity analysis confirmed the findings of the first-to-last comparison: spikes and HFOs showed a consistently strong relationship with the changes in the measured brain dynamics, while drug dosage and other factors did not.

**Figure 5.**
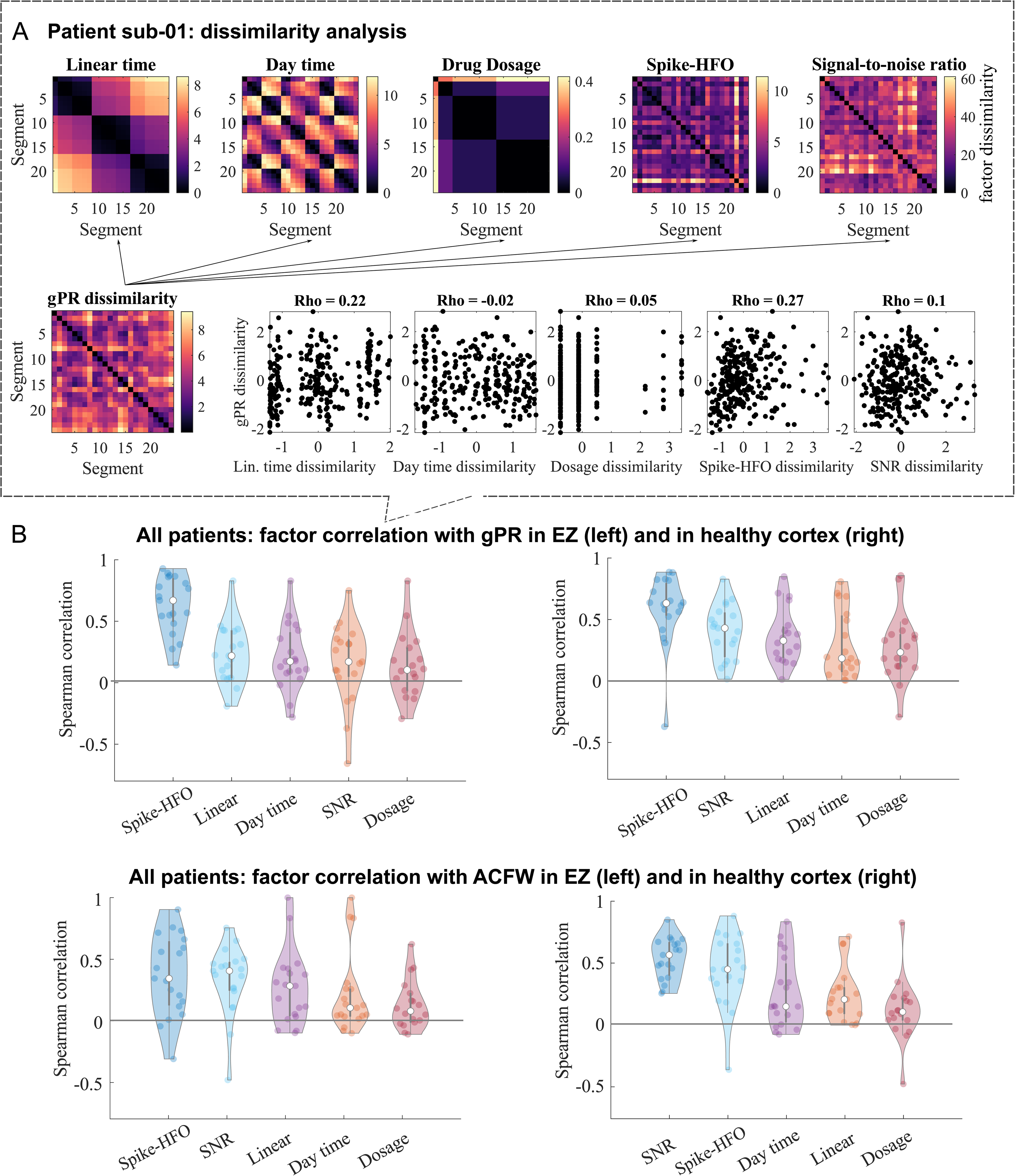
Dissimilarity analysis of factor correlations and the relationship of gPR with spikes and HFOs across patients and zones. As opposed to previous analysis, dissimilarity analysis leverages all available iEEG data, rather than only the first and last segments, estimating the dissimilarity between all possible pairs of segments. A: Dissimilarity in factors (top row) and gPR (bottom left) between all pairs of data segments in exemplary patient Sub-01. For each patient, we then compared the dissimilarity of gPR and factors using Spearman’s rho. Therefore, we obtained a single correlation value for each pair of recordings (bottom right). B: Correlation results for the different factors across patients in EZ and in the healthy cortex. Overall, the relationship between the dissimilarity of measures (gPR and ACFW) and factors (SNR, Spikes, Day time, Linear time, Drug dosage) correlated on average above zero. The factors are plotted in descending order based on the correlation strength. Among the different factors, spikes and HFO rate were most consistently linked to measure changes. Surprisingly, drug dosage had the lowest similarity with measure changes. Spikes in the healthy cortex are most likely due to false detections of the automatic spike detection procedure.

Studying the spike and HFO rate dynamics individually, we observe both increases and decreases across the long-term interval between implantation and the first seizure (Fig. 6). Thus, the link between brain measures and spike and HFO rate dynamics (compare Fig. 1E and Fig. 6A) is more complex than simultaneous increases during the multi-day interval.

**Figure 6.**
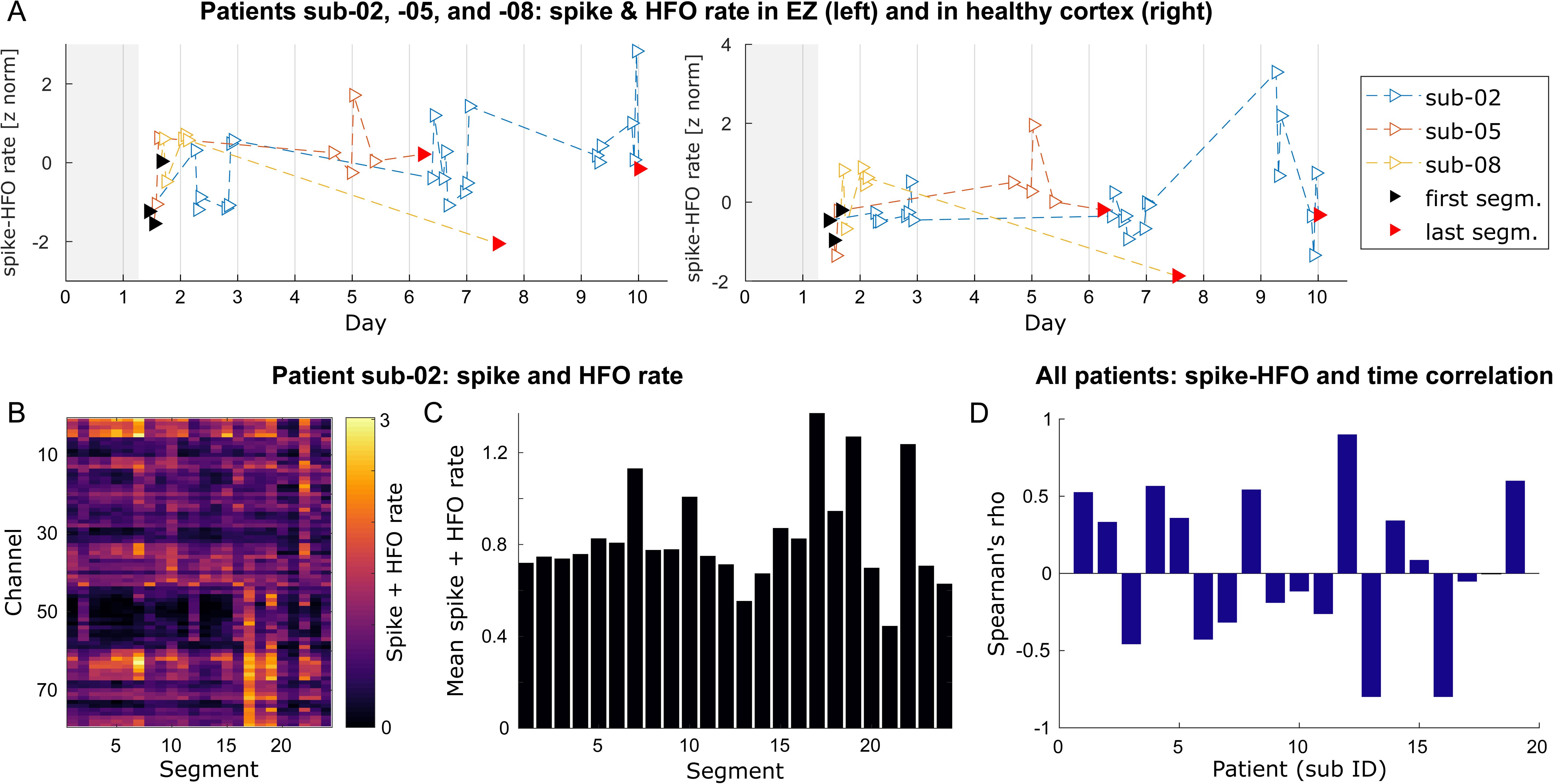
Spike and HFO characteristics in long-term analysis. **A:** Spike and HFO rates as exemplified in three patients: sub-02, sub-05, and sub-08 (see Table 1). Each iEEG segment (triangle) was characterized by a summary of spikes and HFO. **B**: Spike and HFO rate in an exemplary patient (sub-02). We estimated the amount of spike and HFO rate in each channel and each segment. **C**: Average spike and HFO rate. The estimated rates were averaged across channels to provide an estimate for each segment. **D**: Spike and HFO rates over time. Finally, we correlated the segment-specific spike and HFO rates with the times of respective measurements. The resulting correlation varied significantly, suggesting that both increase and decrease in the spike and HFO rates are observable across patients as we approach the seizure.

## 4. DISCUSSION

This study investigated multi-day changes in brain dynamics during the presurgical evaluation of epilepsy patients; more specifically, the time interval between electrode implantation and the occurrence of the first spontaneous seizure ending immediately prior to the clinically marked seizure onset. While we did not find short-term changes within the last 10 minutes before seizure onset (Fig. 2), we find long-term increases in gamma band power prominent in the EZ and a network-wide increase in critical slowing when comparing 10-minute iEEG segments from the beginning and from the end of this multi-day interval (Fig. 4). These findings indicate that the brain dynamic changes during presurgical evaluation could in fact be a multi-day phenomenon with changes occurring on a slow time scale. We found no significant changes in gPLV (short- or long-term), but a potential long-term change could have failed significance testing due to the limited sample size. The identified long-term changes in gPR and ACFW were consistently linked to the long-term changes in number of spikes and HFOs detected. This link was further supported by a dissimilarity analysis (see Fig. 5), exploring the pairwise relationship between all extracted 10-minute segments of iEEG data scattered across the multi-day interval. Other factors, such as drug dose and circadian time did not show a consistent relationship with the identified brain changes. Investigating the long-term spike and HFO dynamics (Fig. 5), we find both patient-specific in- and decreases.

### 4.1. Long-term changes in gamma power and critical slowing before seizure

The novelty of this study was to investigate iEEG changes in the multi-day interval leading up to the first seizure during presurgical monitoring - a time window presumably marked by a progressive increase in seizure likelihood. We studied different time scales (minutes before seizure vs. days before seizure), network locations (EZ vs. healthy cortex) and the interaction with several pro-ictal factors, previously linked to the seizure-modulating processes (Bernard et al., 2023; Karoly et al., 2018; Meisel, 2020; Meisel et al., 2015; Panagiotopoulou et al., 2022; Payne et al., 2021).

Concerning time scales, we observed the changes in gPR and ACFW measures over days, but not over minutes before seizure. The observed long-term increase in gPR and ACFW found in this study confirms previous findings on gamma power increases and critical slowing in the days prior to seizures (Chang et al., 2018; Hughes, 2008; Maturana et al., 2020; Medvedev et al., 2011). They support and extend the findings of Maturana and colleagues (Maturana et al., 2020), who showed that seizures align with the increasing phase of ACFW on a slow time scale. Moreover, slow fluctuations in iEEG power as described in Panagiotopoulou and colleagues (Panagiotopoulou et al., 2022) could be linked to the multi-day gPR changes identified in this study, but not to circadian fluctuations as we did not find consistent correlations with the factor of daytime. We interpret the absence of a short-term increase pre seizure in these measures such that the observed changes capture pro-ictal brain states (during an increased seizure likelihood), as opposed to pre-ictal states (during a single seizure transition; see detailed discussion below).

For network locations, gPR increased locally in EZ (same as in (Medvedev et al., 2011)). Aberrant dynamics in epilepsy are commonly found within the epileptogenic network (Bartolomei et al., 2017). However, other studies have reported changes outside the epileptogenic network, complementary to those inside (Naftulin et al., 2018). Unlike in Naftulin and colleagues we did not see indications of a systematic decrease in gamma power outside of the epileptogenic network (healthy cortex, see Fig. 3). ACFW on the other hand showed a network-wide increase, supporting Maturana and colleagues’s hypothesis, that critical slowing characterizes a state change throughout the whole brain, rather than in a localized brain region (Maturana et al., 2020). Please note that the presented analyses were conducted using averages across EZ and healthy cortex. Future studies could refine this approach by analyzing individual iEEG channels, for instance, by identifying distinct patterns of epileptogenic activity through clustering analysis of channel-specific temporal trajectories. Additionally, applying directed connectivity measures could help uncover the directionality of interactions between individual channels. Channel-resolved analyses could further investigate the significant correlation of long-term change in ACFW and the number of channels in EZ, as identified in the post-hoc analysis, which could hint at a potential influence of electrode placement variations across patients in this study.

For the studied pro-ictal factors, spikes and HFOs, considered hallmarks of epilepsy (Zijlmans et al., 2019), showed a consistently significant link to the multi-day changes in autocorrelation and critical slowing. Other factors, including linear, circadian time and drug dose did not show a consistent link with the observed measure changes in this study. The relationship between pre-ictal brain dynamics and spikes and HFOs was previously established (Jacobs et al., 2009; Malinowska et al., 2015; Scott et al., 2021), especially between spikes and HFOs and the changes in high-frequency spectral power (Panagiotopoulou et al., 2022) and the measures of critical slowing (Maturana et al., 2020). Importantly, gamma power changes remain after removing spikes from the iEEG signal via signal processing (Jmail et al., 2017). Thus, spikes and HFOs and gamma power can be assumed to capture complementary signal characteristics. It is important to note that, in our study, the maximum frequency of HFO assessment was limited to 126 Hz due to the sampling frequency of the data, and that there was a partial overlap of the frequency ranges of the two (gPR: 30-100 Hz; HFOs: 80-126 Hz).

A major limitation to this study was the relatively low amount of data (29 patients in the short-term analysis with one 10-minute segment per patient, and 19 patients in the long-term analysis with two 10-minute segments each, including the short-term segment). Additionally, for the explorative part of the long-term analysis, each patient had an average of 20 iEEG segments, spread across a maximum of 12 days. The irregular data sampling did not allow us to statistically investigate the multi-day progression of brain measure and factor changes. A continuous dataset is needed to allow the study of the detailed progression of brain states during this interval of presurgical evaluation.

Nevertheless, our results support the potential usefulness of the measures studied to serve as markers of seizure-modulating processes (Maturana et al., 2020; Panagiotopoulou et al., 2022). Serving as such, if implemented in clinical practice, they could help improve the yield of the pre-surgical evaluation, which is a long and cumbersome procedure; e.g. help to reduce recording length, identifying time widows for enhanced surveillance of patients in a hospital video-EEG unit, choose the best time window for stimulation probing, or optimize the drug reduction protocol to avoid withdrawal complications. An advantage of the measures used in this study is that they are easily applicable in an automatized fashion and computationally inexpensive. As such, they could also be incorporated into ambulatory devices. Correlating the pro-ictal markers with the patient’s environment would allow for better identification of precipitating factors and adjustments to a patient’s daily life in order to reduce the seizure risk. However, assessing the practical application of pro-ictal markers remains the task of future studies.

### 4.2. Pre-ictal and pro-ictal brain states and the origin of long-term fluctuations in seizure susceptibility

The distinction of a so-called *pro-ictal* brain state from previously studied pre-ictal states is suggested by several recent studies evidencing the existence of time windows of an increased seizure risk and slow fluctuations in seizure susceptibility over time (see (Baud et al., 2020) for a recent review). The results of the current study contribute to the open discussion regarding the origin of these fluctuations and the relationship between pre- and pro-ictal states in epilepsy. While approaches to track pre-ictal and pro-ictal states are clearly intertwined, the underlying brain processes they target operate on rather different timescales. These processes may therefore be inherently different in nature and subject to distinct modulation by external factors (e.g. a flashing light might cause a single seizure transition but not lead to a more long-term increase in seizure likelihood, or sleep and wake cycles might modulate the rate of seizure occurrence but not change the characteristics of the individual seizure transitions). Moreover, an interplay of the two is possible, where pre-ictal dynamics are superimposed on pro-ictal dynamics. An analogical example for such a superposition could be the occurrence of lightning during thunderstorms, where certain weather conditions (“*pro*-lightening” states) are necessary for enabling the lightning build-up-and-discharge mechanisms (including “*pre*-lightening” states). Understanding the (potentially patient-specific) interplay of pro-ictal and pre-ictal states remains an open challenge.

In this study, we adopted tools from seizure prediction and forecasting aimed at identifying pre-ictal and pro-ictal brain states, in an effort to help disentangle the complex mechanisms involved in seizure generation. The chosen multi-day dataset of presurgical iEEG monitoring in epilepsy patients, even though it was comparably small and did not provide continuous or regularly sampled recordings, provided the means to compare the short-term (minutes before a seizure) and long-term (days before a seizure) time scales. While we found brain dynamic changes in the long-term, we did not find changes in the short-term. Importantly, in the studied dataset, seizure susceptibility mechanisms were manipulated by decreasing the administered drug dose (Hartl et al., 2019). Thus, a novelty of this study was to apply measures from seizure prediction in the context of a controlled progressive loss of seizure resilience (Chang et al., 2018) that should give rise to a pro-ictal state. We argue that the identified differences over days, but not minutes, are likely to reflect brain changes characteristic of pro-ictal states as opposed to pre-ictal states of individual seizure transitions. As in the case of pre-ictal states, an observed build-up should continue until seizure is reached. The missing short-term increase in this study could be an indication that the detected brain changes do, in fact, reflect a pro-ictal state. We did not, however, find the expected correlation with linear time and drug dosage decrease, even though previous accounts have linked anti-seizure medication effects and critical slowing and high-frequency changes (Duncan, 1987; Lepeu et al., 2024; Meisel, 2020). The identified significant correlation of pro-ictal brain changes and the measured rate of spikes and HFOs is in line with previous literature on pro-ictal states (Baud et al., 2020) and their considered important role in epilepsy diagnosis (Zijlmans et al., 2019). The tentative causal mechanisms involved in their interaction with seizures remain a field of on-going research (Jiruska et al., 2017).

## 5. CONCLUSIONS

This study highlights gamma oscillations and critical slowing as markers of pro-ictal, multi-day brain dynamics during the presurgical evaluation of epilepsy patients. While gamma power increased locally in EZ, critical slowing increased network-wide across the different regions. Interestingly, the rate of spikes and HFOs, a hallmark of epilepsy, was consistently linked with the significant changes in brain dynamics identified in this study. Thus, gamma power and critical slowing could be useful tools to track epileptic brain states for the study of epilepsy mechanisms, or in the clinical or ambulatory setting, to track changes in seizure susceptibility over days. We encourage future studies in larger and continuously recorded cohorts, to better understand pro-ictal brain dynamics and to assess the clinical usefulness of pro-ictal state assessment.

## Data Availability

Data available on request due to privacy/ethical restrictions.

## DECLARATIONS

## Ethical publication statement

We confirm that we have read the Journal’s position on issues involved in ethical publication and affirm that this report is consistent with those guidelines.

## IRB statement

The ethical conditions of this study have been upheld and approved by the Toulouse University Hospital, where the study is registered in the official retrospective studies register, managed by the Research and Innovation Department (RnIPH 2020-130), and is covered by the MR-004 law (CNIL number: 22.6723 v 0). Consent was obtained from all patients.

## Credit authorship contribution statement

IDZ, JK, AP, JC, EJB and JH conceived the study. JC, MD, JCS and LV are responsible for data collection and curation. IDZ, JK and AP analyzed the data under supervision of JH. IDZ and JK wrote the manuscript with contributions from AP, JC, EJB and JH. All co authors reviewed the manuscript.

## Acknowledgement

This work was supported by the bilateral Barrande Mobility scheme, funded by the Ministry of Education, Youth and Sports of Czech Republic, project number MSMT–7125/2020-16 8J20FR037 and Campus France grant No. 44815QG, the Czech Science Foundation. project No. 21-32608S, by ERDF-Project Brain dynamics, No. CZ.02.01.01/00/22_008/0004643, and the long-term strategic development financing of the Institute of Computer Science (RVO:67985807) of the Czech Academy of Sciences.

## Availability of data and materials

The datasets generated and/or analyzed during the current study are not publicly available due privacy/ethical restrictions but are available from the corresponding author on reasonable request.

