## Supplementary Material for "Pro-ictal, rather than pre-ictal, brain state marked by global critical slowing and local gamma power increase"

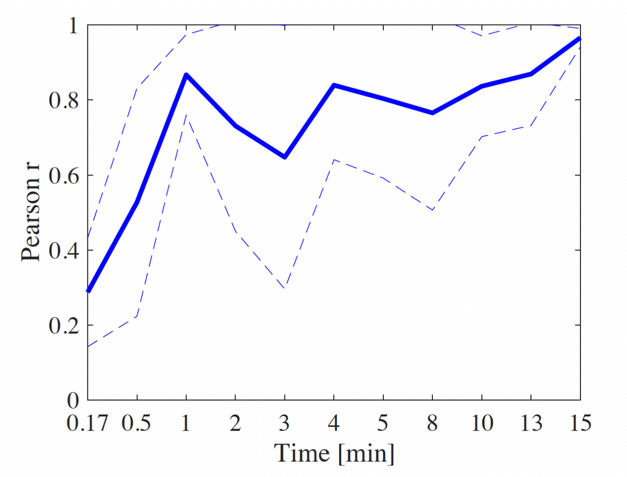


### Supplementary Figure 1

***Selection of analysis segment length.*** *The selection of optimal segment length was based on the stability of gPLV. We first estimated gPLV in two-hour-long uninterrupted iEEG recording from a healthy cortex. In the next step, we estimate gPLV in 10,000 shorter iEEG segments of varying lengths, ranging from 10 seconds to 15 minutes, randomly sampled from the original two-hour-long iEEG segment. The random sampling approach ensures that the analysis captures a diverse range of iEEG activity, making the stability assessment robust. Finally, we quantified the correlation between gPLV values from the original 2-hour recording and those from the shorter segments. The figure depicts the mean ± standard deviation of Pearson's correlation for each segment size. Ultimately, 10-minute segments were selected as heuristically balancing sufficient length for stable gPLV estimates with a large number of available segments for further analysis.*

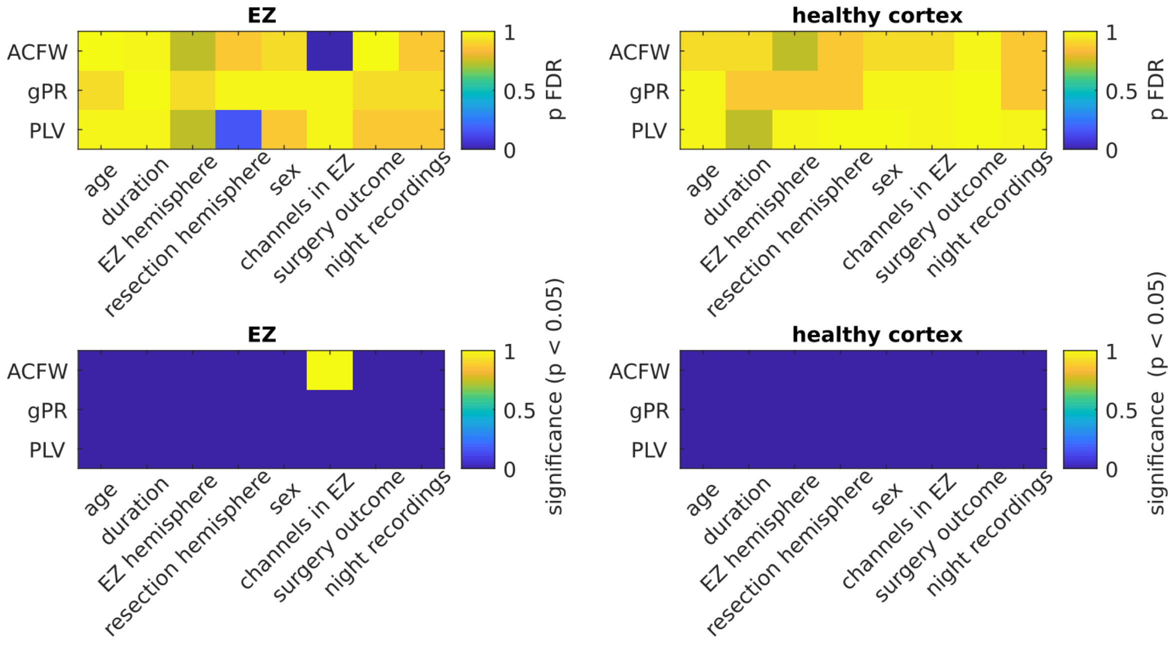


### Supplementary Figure 2

***The association of ACFW, gPR, and gMPC with patient variables.*** *We tested for the group difference in the ACFW, gPR, and gMPC change based on surgery outcome (ILAE 1 vs. ILAE 3-5), sex (male vs. female), and hemisphere of EZ and resection (left vs. right). In addition, we compared ILAE 1 against ILAE 3-5. The p-value of Spearman's correlation was used to determine the significance. Furthermore, we extended this analysis to other qualitative clinical variables (number of electrodes, number of channels, time since epilepsy diagnosis at the time of hospitalization, and number of electrodes, number of recordings per night). Resulting p-values were corrected for multiple comparisons using FDR correction across measures and zones. Only one of the linear associations exceeded the significance level, namely the change in ACFW in EZ and the numbers of electrodes in EZ.*

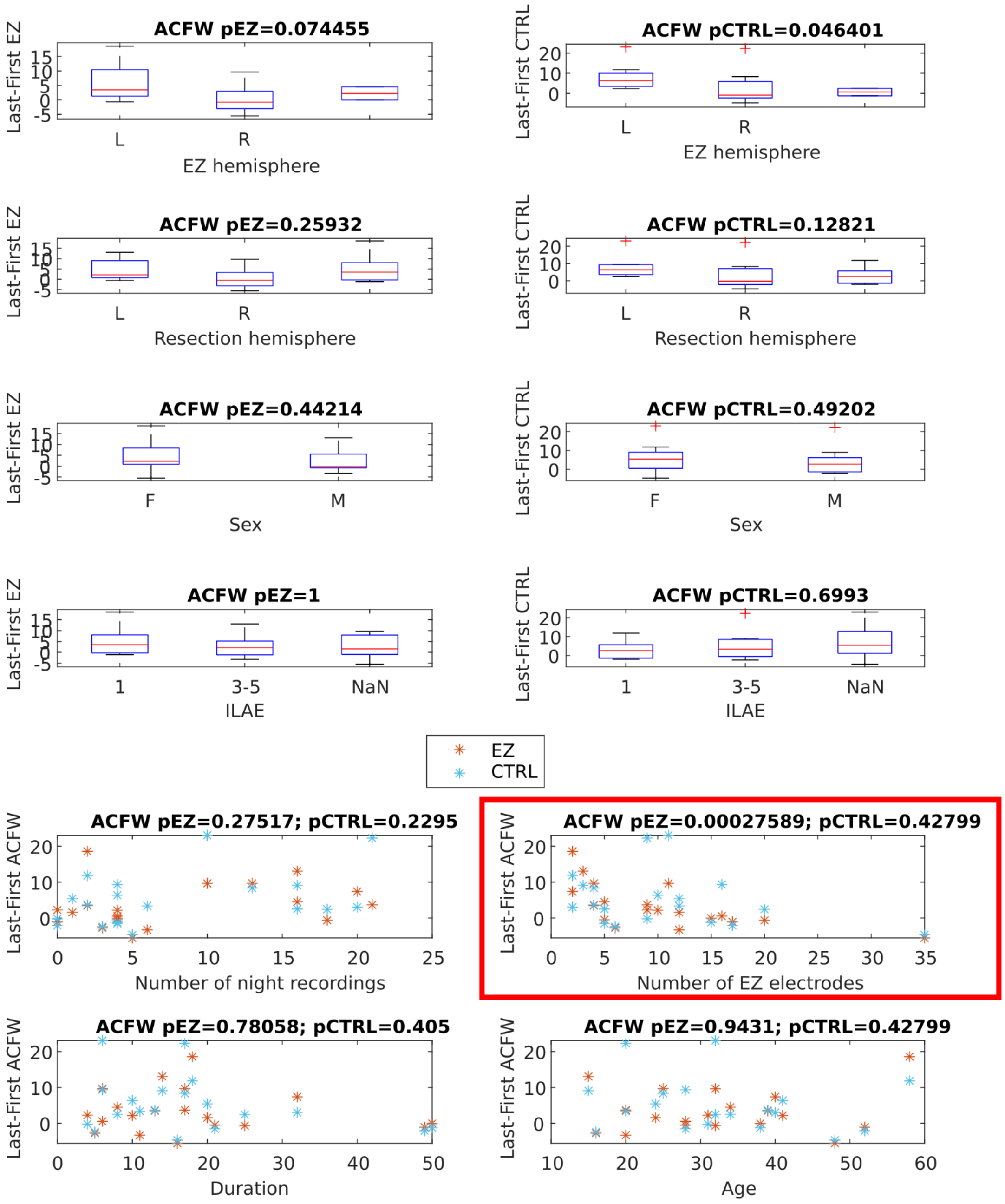


### Supplementary Figure 3

***The association of ACFW with patient variables.*** *Note that the depicted p-values are before FDR correction. For FDR corrected values see previous figure. There was a significant association between the numbers of channels localized in EZ and ACFW change from first to last, marked here in red frame. The scatter plots depict continuous variables compared to measure change, for EZ (in blue) and healthy cortex (orange). p values mark the significance of Spearman's correlation. Boxplots in the panels above display relationships between measure changes and categorical variables, surgery* *outcome (ILAE 1 vs. ILAE 3-5), sex (male vs. female), and hemisphere of EZ and resection (left vs. right), tested using Wilcoxon rank sum test. EZ: epileptogenic zone, CTRL: healthy cortex.*

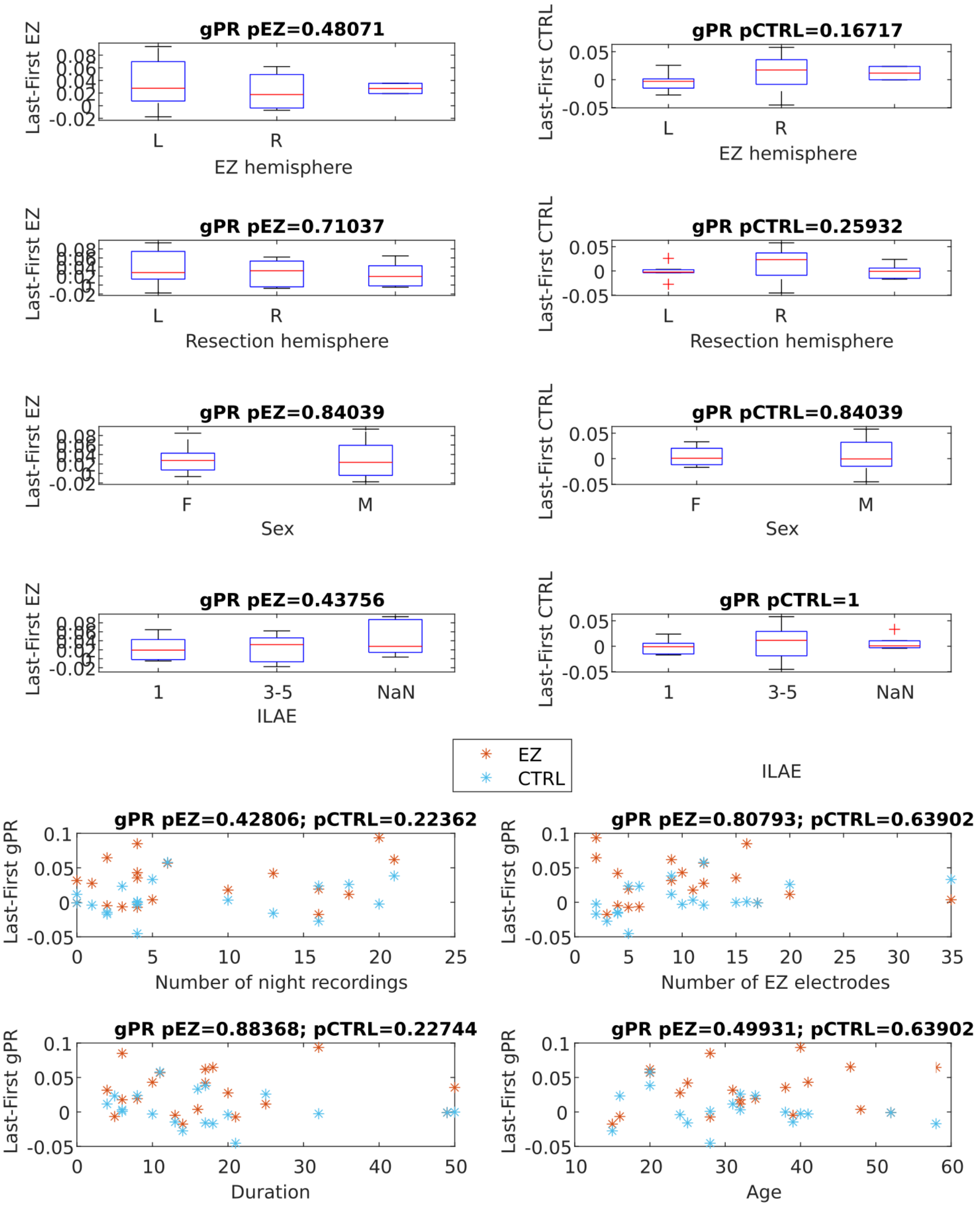


### Supplementary Figure 4

***The association of gPR with patient variables.****. We did not find any significant association or difference between any variable and gPR change (see previous captions for figure description).*

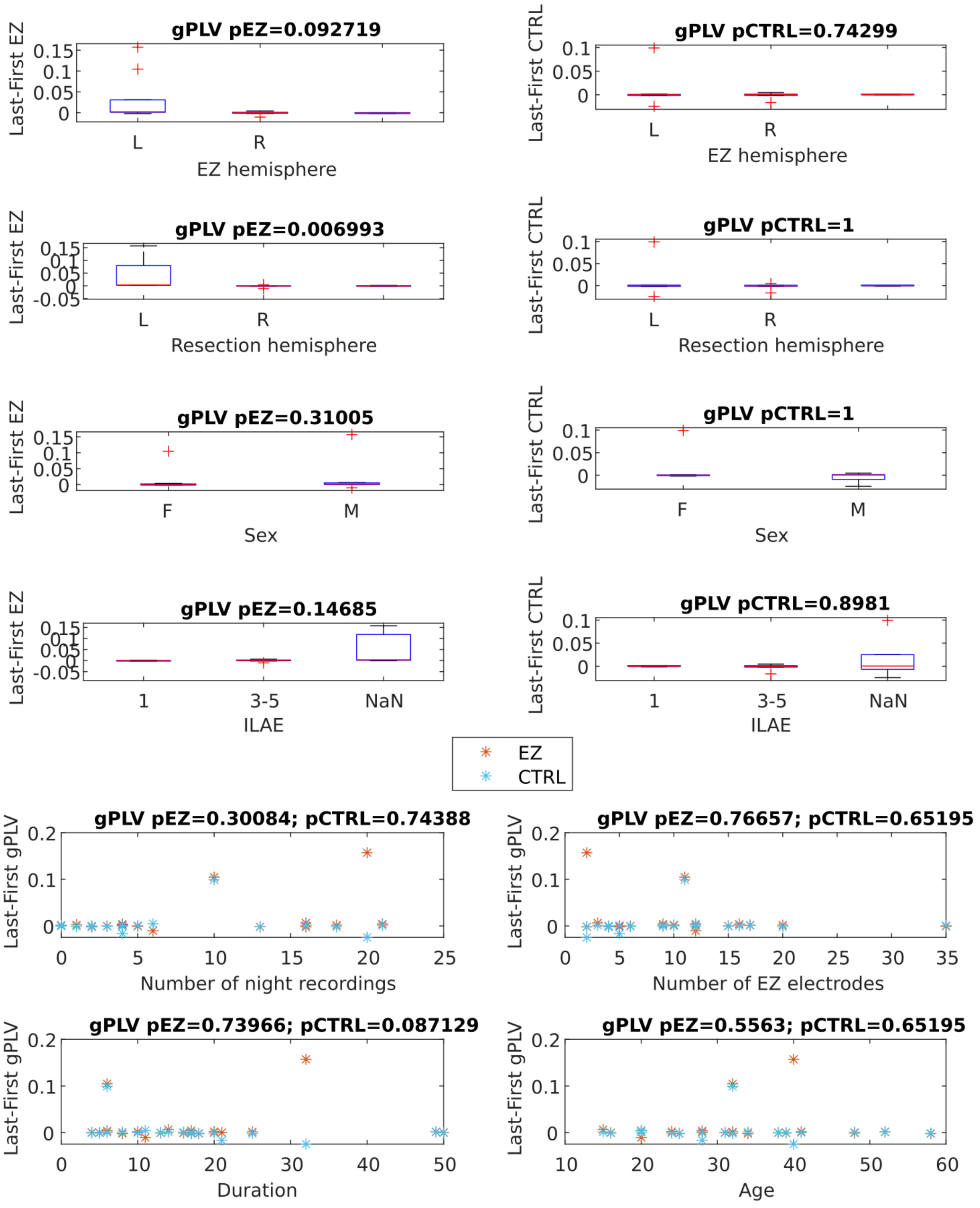


### Supplementary Figure 5

***The association of gPLV.*** *After FDR correction, we did not find any significant association or difference between any variable and gPLV change (see previous captions for figure description).*

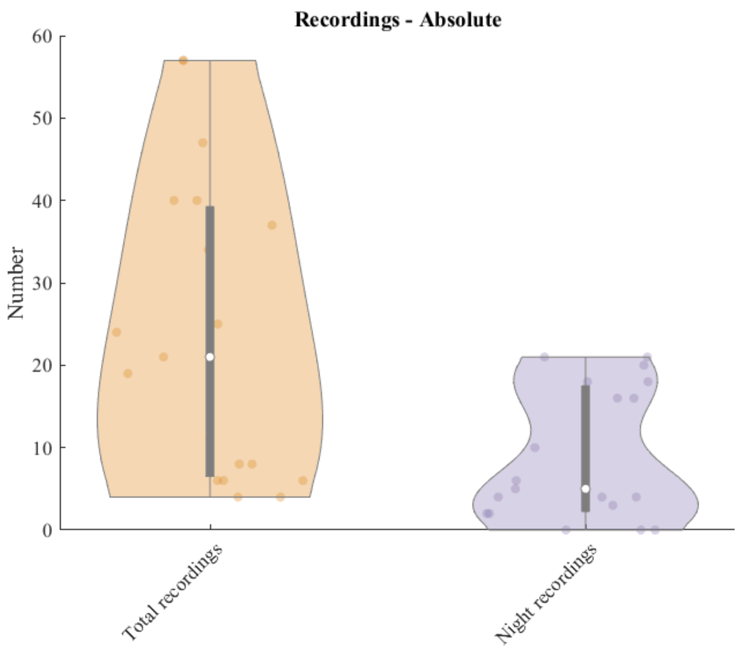


### Supplementary Figure 6

***Number of recordings (total and per night) used in the long-term analysis.*** *Each dot is a patient. Sleep and awake states were not tracked in this study. The numbers of recordings during nights (defined as recording start time between 21:59 and 6:59) could suggest a potential influence of arousal states to our main finding. However, significance testing of a relationship between the number of night recordings and the change in ACFW, gPR, and gMPC did not yield significant results (see previous Supplementary Figures).*
